# Higher COVID-19 Vaccination Rates among Unemployed in the United States: State Level Study in the First 100 days of Vaccine Initiation

**DOI:** 10.1101/2021.04.17.21255668

**Authors:** Ali Roghani, Samin Panahi

## Abstract

**Background:** Socioeconomic factors may impact the efficiency of COVID vaccine rollout; however, there are limited studies that examine how state socioeconomic status influences the speed of vaccine distribution. This study aimed to demonstrate how employment rates as one of the main socioeconomic factors affect vaccine coverage in 50 states of the United States.

**Methods:** This study has obtained vaccine data for the 50 states in the United States, available in the electronic online database ourworldindata.org. In addition to employment rates, other socioeconomic determinants including poverty level, uninsured rates, population density, homeownership, educational level, the percentage of the elderly population, and educational level were obtained from ourworldindata.org data platform. Data from these 50 states were used for regression analyses to examine the relationship between socioeconomic and vaccination rates.

**Results:** Our study revealed a positive linear association between unemployment and vaccine rates, and states with higher unemployment rates were more likely to have higher vaccination rates. However, other socioeconomic measures do not significantly associate with vaccine coverage.

**Conclusion:** Despite other studies showing that vulnerable populations had lower vaccine rates, this study shows that states with higher unemployment rates are more likely to be vaccinated. However, the finding suggests a need for more research for the states with higher than 5% unemployment rates, as they had a lower vaccine coverage than states with a range of 4% to 5% unemployment rates.

## Introduction

The United States has been one of the fastest countries in COVID-19 vaccine rollout globally (1). However, many factors may influence the efficiency and speed of vaccine rollout, which lagged some states considerably behind the average. Although it is well known that socioeconomic (SES) determinants have a tremendous impact on a faster vaccine rate globally (2), there are limited studies that examine the relationship between SES and vaccine level at a state level. Previous studies show that COVID-19 has impacted those economically disadvantaged (3); therefore, achieving vaccine equality and equity are crucial factors that can protect more vulnerable populations.

Previous research indicates COVID-19 vaccination equity differed among states in which coverage was greater in low vulnerability counties than in high vulnerability counties. However, states such as Arizona and Montana had higher vaccination rates in high vulnerability counties (3). Lindemer and colleagues showed counties with high uninsured rates and unemployed adults have lower vaccination rates (4). Therefore, it can be concluded lower socioeconomic status is associated with a lower vaccine. Using publicly available data, we evaluate the association between unemployment rates and vaccine coverage in 50 states across the United States. This study will demonstrate that variables related to socioeconomic status predict vaccine coverage.

## Methods

We analyzed 50 states in the United States that have cumulative vaccination data available up to April 2021. Ninety-nine million and five hundred sixty-five thousand and three hundred individuals received at least one dose, and 56,089,614 were fully vaccinated. Vaccination data were obtained from https://ourworldindata.org/covid-vaccinations(5). For each state, the vaccination rate was defined as the percentage of individuals in the county with at least one dose of an FDA-emergency authorized COVID-19 vaccine. Figure 1 and 2 show vaccine distribution at the end of March, which indicate 60% residents of some states received the first dose and in some states, 20% of residents were vaccinated. Demographic and socioeconomic data for each state were obtained from the https://github.com/stccenter/COVID-19-Data/tree/master/US. The leading independent variable is state-level unemployment claims, which can be used in any economic analysis of unemployment trends in the United States. Initial claims measure emerging unemployment, and continued weeks count the number of persons claiming unemployment benefits. The data sheds light on the current economic response to COVID-19 and how the numbers in certain states fluctuate over time. This research used the first-week unemployment rates in March to examine the impact of unemployment on COVID-19 vaccine distribution. To determine the association between unemployment rate and vaccine rates, ordinary least squares regression was used. Model 1 is the regression of unemployment rates and vaccine distribution, and Model 2 is adjusted, including other socioeconomic and demographic factors.

**Figure 1.**
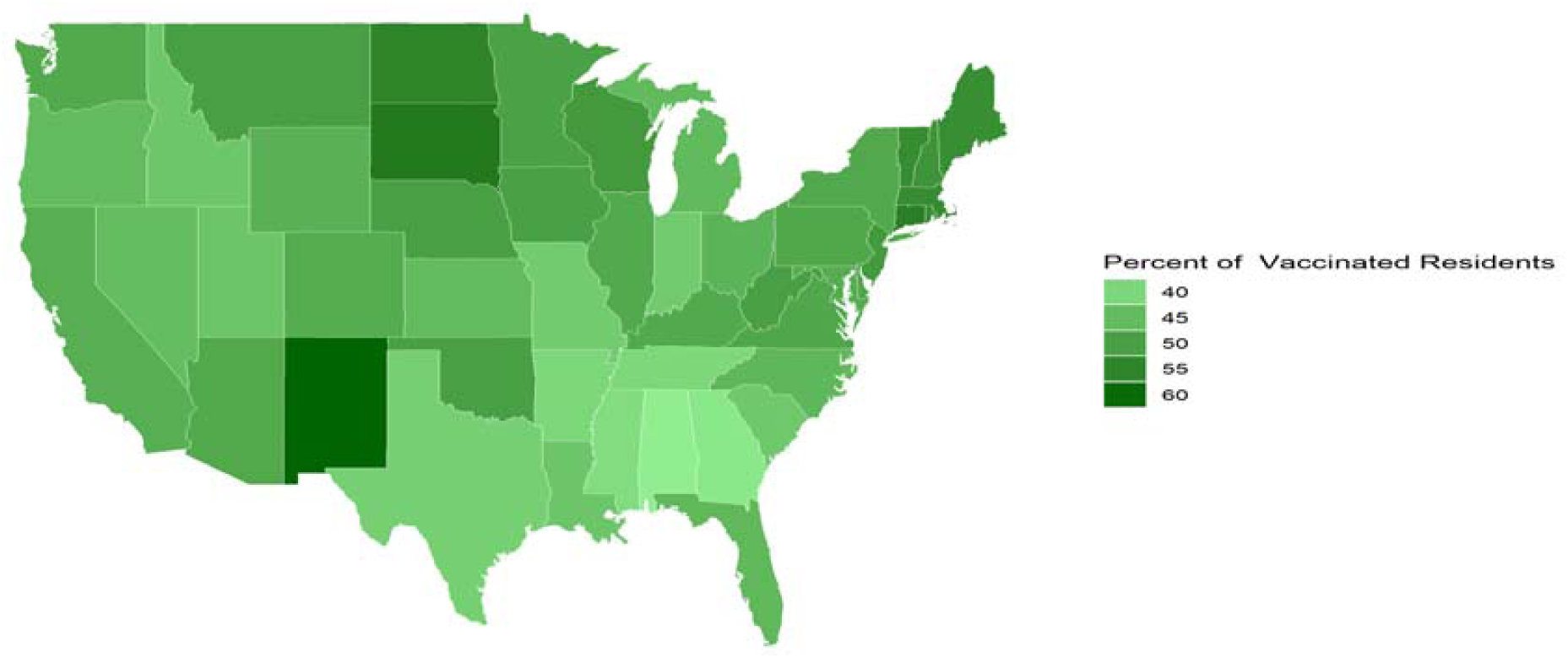
Percent of Delivered First Vaccine Doses by State in the US.

**Figure 2.**
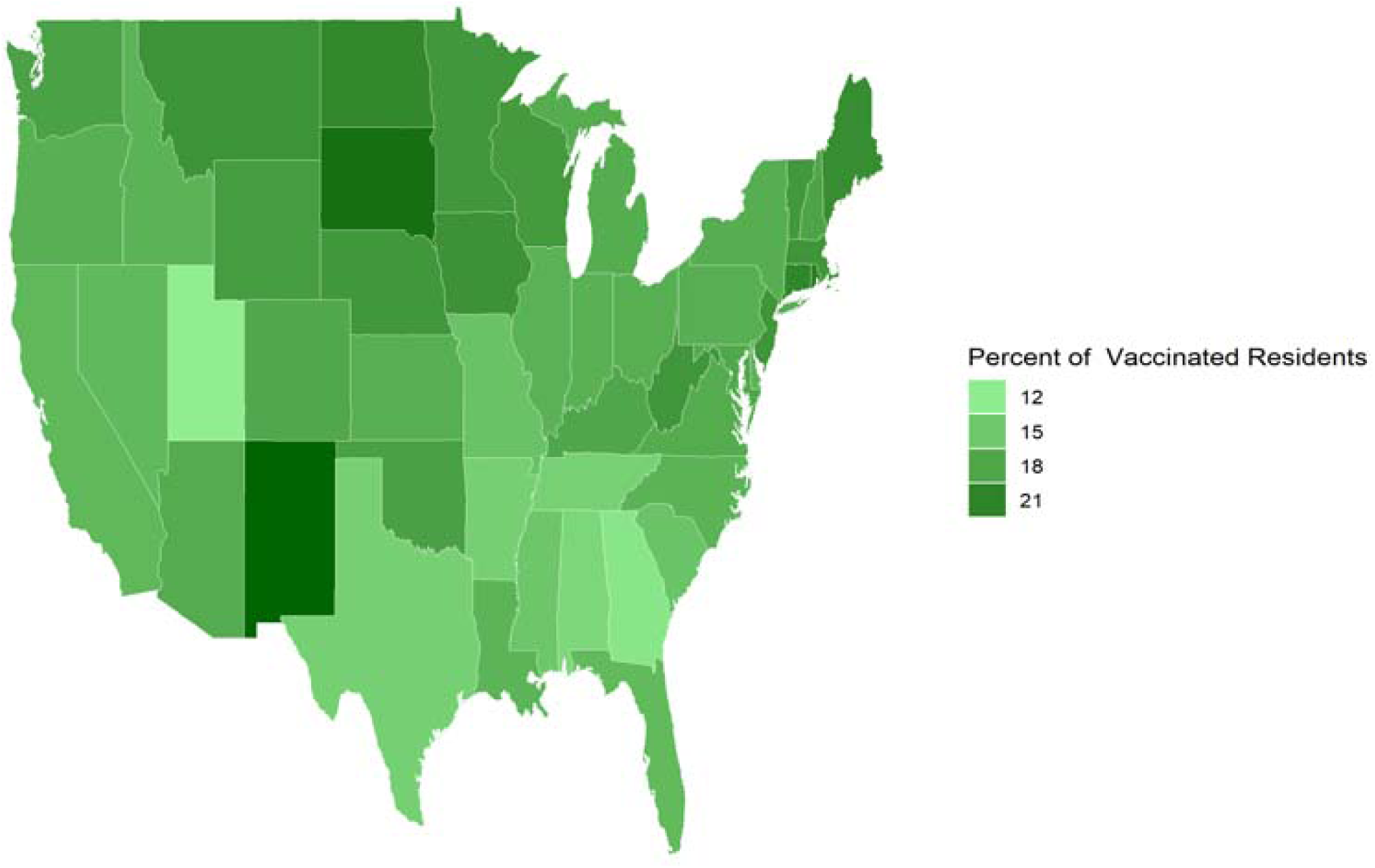
Percent of Fully Vaccinated Residents by State in the US.

## Result

Alaska, Connecticut, and Nevada have the highest unemployment rates (more than 5%) in the US in March 2021. North Dakota, New Mexico, Wisconsin, and Minnesota have the highest percentage of COVID-19 vaccine rates. In terms of other demographic determinants, Texas has the highest rate of uninsured residents. Mississippi and Las Vegas have the highest poverty rate in the US. Maine is the oldest state in the US, and New Jersey has the highest percentage of the White population. Wyoming residents have the highest rate of homeownership, and Colorado residents are the most educated in the US.

Table 2 shows the relationships between the measure of the unemployment rate and vaccine distribution. The scatterplot also indicates a positive linear relationship between the unemployment rate and vaccine distribution. The range of unemployment rate between states was from 0.90% to 4.69%. The regression coefficient indicates that a 1 unit increase in unemployment rate coefficient was associated with a rise of 1.6 vaccine distribution (p <0.001). Although the association has been positive and linear, states with higher than 5 percent had lower vaccine distribution than their counterparts between 4 to 5 unemployment rates. The second model is adjusted analysis, which shows higher unemployment is associated with higher vaccine coverage, and states with higher population density are less likely to have higher vaccine distribution. Higher vaccine coverage is across all states with higher unemployment rates; however, this relationship is not consistent in states whit higher than 5% unemployment rates.

**Table 1.**

| Postal Code | Insured Unemployment Rate | Uninsured | Poverty rate | Population size | Senior Population | White Population | House Owner | Bachelor degree |
| --- | --- | --- | --- | --- | --- | --- | --- | --- |
| AL | 6.00 | 9.7 | 15.5 | 4803185 | 17.4 | 65.5 | 68.8 | 16.3 |
| AK | 5.58 | 12.2 | 10.1 | 731545 | 12.4 | 71.4 | 84.7 | 18.5 |
| AZ | 2.09 | 11.3 | 13.5 | 7279717 | 18.0 | 81.7 | 65.3 | 18.8 |
| AR | 2.11 | 9.1 | 18.2 | 3017804 | 17.4 | 79.3 | 65.5 | 15.1 |
| CA | 4.16 | 7.7 | 11.8 | 39512223 | 14.8 | 63.6 | 54.9 | 21.9 |
| CO | 3.49 | 8.0 | 9.3 | 5759736 | 14.7 | 87.3 | 65.9 | 26.6 |
| CT | 5.01 | 6.6 | 10.6 | 3666287 | 17.6 | 77.8 | 66.0 | 22.0 |
| DE | 2.77 | 6.6 | 11.3 | 973784 | 19.5 | 70.5 | 70.3 | 19.5 |
| DC | 3.46 | 3.5 | 13.5 | 705749 | 12.4 | 45.1 | 41.5 | 25.7 |
| FL | 1.66 | 13.2 | 12.7 | 21477737 | 20.9 | 77.1 | 66.2 | 10.3 |
| GA | 3.26 | 13.4 | 13.3 | 10617423 | 14.3 | 58.9 | 64.1 | 19.9 |
| HI | 3.15 | 4.2 | 9.3 | 1415872 | 19.0 | 41.1 | 60.2 | 22.1 |
| ID | 1.90 | 10.6 | 11.2 | 1707005 | 16.2 | 92.5 | 71.0 | 18.8 |
| IL | 4.35 | 7.4 | 11.5 | 12671821 | 16.1 | 73.8 | 66.0 | 21.7 |
| IN | 2.35 | 8.7 | 11.9 | 6732219 | 16.1 | 65.2 | 69.2 | 17.3 |
| IA | 2.94 | 5.3 | 11.2 | 3155070 | 17.5 | 91.9 | 70.5 | 19.8 |
| KS | 1.86 | 9.2 | 11.4 | 2913314 | 16.4 | 87.1 | 66.5 | 21.6 |
| KY | 2.43 | 6.4 | 16.3 | 4467673 | 16.9 | 88.9 | 67.0 | 14.9 |
| LA | 2.83 | 8.9 | 19.0 | 4648791 | 16.0 | 63.5 | 66.5 | 16.0 |
| ME | 2.79 | 8.0 | 10.9 | 1344212 | 21.3 | 95.9 | 72.2 | 20.8 |
| MD | 2.54 | 6.0 | 9.0 | 6045680 | 15.9 | 57.3 | 66.6 | 21.8 |
| MA | 4.36 | 3.0 | 9.4 | 6852503 | 17.0 | 80.2 | 62.2 | 24.7 |
| MI | 3.86 | 5.8 | 13.0 | 9966857 | 17.7 | 81.0 | 71.6 | 18.2 |
| MN | 3.99 | 4.9 | 9.0 | 5659632 | 16.3 | 85.1 | 71.5 | 24.5 |
| MS | 2.80 | 13.0 | 19.0 | 2970149 | 16.4 | 59.4 | 67.3 | 13.7 |
| MO | 2.99 | 10.0 | 12.9 | 6137423 | 17.2 | 84.5 | 67.1 | 18.4 |
| MT | 3.00 | 8.3 | 12.6 | 1068773 | 19.5 | 91.1 | 68.5 | 23.1 |
| NE | 1.48 | 8.3 | 9.9 | 1934403 | 16.1 | 86.7 | 66.2 | 21.8 |
| NV | 5.43 | 11.4 | 12.5 | 3060150 | 16.2 | 68.4 | 56.0 | 16.7 |
| NH | 3.93 | 6.3 | 7.3 | 1350711 | 18.8 | 94.7 | 71.0 | 22.9 |
| NJ | 3.29 | 7.9 | 9.2 | 8852190 | 16.5 | 69.5 | 63.3 | 25.1 |
| NM | 4.18 | 10.0 | 16.2 | 2060629 | 18.0 | 76.6 | 68.1 | 15.5 |
| NC | 1.37 | 11.3 | 13.6 | 10488081 | 16.7 | 70.5 | 66.3 | 20.5 |
| ND | 2.27 | 6.9 | 10.6 | 762062 | 15.8 | 89.0 | 61.3 | 21.5 |
| OH | 3.41 | 6.5 | 13.1 | 11069100 | 17.5 | 83.5 | 66.0 | 18.2 |
| OK | 2.25 | 14.3 | 15.2 | 3956971 | 16.1 | 79.2 | 65.5 | 17.1 |
| OR | 3.70 | 7.2 | 11.4 | 4217737 | 18.2 | 88.1 | 62.9 | 21.0 |
| PA | 6.35 | 5.5 | 12.0 | 12801989 | 18.7 | 81.9 | 68.4 | 19.5 |
| RI | 4.54 | 4.1 | 10.0 | 1059301 | 17.7 | 62.0 | 61.7 | 20.9 |
| SC | 2.95 | 10.9 | 19.8 | 5148711 | 18.2 | 68.8 | 70.3 | 18.4 |
| SD | 1.38 | 10.2 | 11.9 | 864659 | 17.4 | 86.7 | 67.6 | 20.6 |
| TN | 1.53 | 10.1 | 13.9 | 6829174 | 16.7 | 79.3 | 66.0 | 16.0 |
| TX | 2.59 | 18.4 | 13.6 | 28965881 | 12.9 | 75.9 | 61.6 | 20.0 |
| UT | 1.03 | 9.7 | 8.9 | 3265958 | 11.4 | 90.2 | 73.6 | 23.4 |
| VT | 4.21 | 4.5 | 10.2 | 623989 | 20.1 | 96.2 | 73.9 | 22.7 |
| VA | 1.79 | 7.9 | 9.9 | 6535519 | 15.9 | 70.2 | 63.1 | 22.4 |
| WA | 3.92 | 6.6 | 9.8 | 7614393 | 15.9 | 79.5 | 63.1 | 22.8 |
| WV | 3.35 | 6.7 | 16.0 | 1792147 | 20.5 | 94.7 | 73.4 | 12.6 |
| WI | 3.61 | 5.7 | 10.4 | 5822434 | 17.5 | 67.4 | 67.2 | 20.7 |
| WY | 2.01 | 12.3 | 10.1 | 578759 | 17.1 | 63.1 | 71.9 | 18.8 |

**Table 2.**

| *Predictors* | **Model 1** |  |  | **Model 2** |  |  |
| --- | --- | --- | --- | --- | --- | --- |
|  | *Estimates* | *CI* | *p* | *Estimates* | *CI* | *p* |
| Intercept | 37.63 | 34.22 – 41.04 | **<0.001** | 38.16 | -0.17 – 76.48 | 0.051 |
| Unemployment Rate | 1.68 | 0.63 – 2.73 | **0.002** | 1.41 | 0.29 – 2.53 | **0.014** |
| Uninsured |  |  |  | 0.01 | -0.51 – 0.53 | 0.970 |
| Poverty Rate |  |  |  | -0.09 | -0.90 – 0.73 | 0.831 |
| Population Size |  |  |  | -0.00 | -0.00 – -0.00 | **0.024** |
| Senior Population |  |  |  | 0.48 | -0.26 – 1.21 | 0.196 |
| White Population |  |  |  | 0.08 | -0.05 – 0.20 | 0.214 |
| House Owner |  |  |  | -0.25 | -0.60 – 0.10 | 0.158 |
| Bachelor Degrees |  |  |  | 0.24 | -0.51 – 0.99 | 0.520 |
| Observations | 50 |  |  | 50 |  |  |
| R<sup>2</sup> / R<sup>2</sup> adjusted | 0.178 / 0.161 |  |  | 0.390 / 0.271 |  |  |

## Discussion

This study examines how the unemployment rate as one of the leading indicators of socioeconomic status predicts vaccine distribution at the USA’s state level. Despite previous studies indicating a strong positive correlation between low levels of health insurance coverage and low vaccination rates(3, 4), the current study’s findings show higher unemployment is associated with higher vaccination rates. The present study examines other socioeconomic factors, including poverty level, uninsured rates, population density, homeownership, educational level, the percentage of the elderly population, and educational level. Unlike higher population density which is associated with lower vaccination rates, other socioeconomic measures do not significantly associate with vaccine coverage.

One reason that can explain the opposite direction between unemployment and vaccine rates can be the vaccine eligibility of unemployed people, and they might have enough knowledge about vaccine eligibility from their primary care providers. This association does not exist for other socioeconomic and demographic factors. The findings highlight the complexities of socioeconomic disparities in the United States, while the USA’s vaccine distribution policy has successfully reduced inequalities despite higher unemployment rates. This study shows improved public health, emphasizing that employment is unnecessary for any of the FDA-authorized COVID-19 vaccines in the United States. However, figure 3 indicates that states with higher than 5% percent did not have similar outcomes than other states that have a range of 4% to 5% unemployment rates. This can imply that although higher employment is associated with higher vaccination rates in the US, this association might not be consistent in the states with the highest unemployment rates.

**Figure 3.**
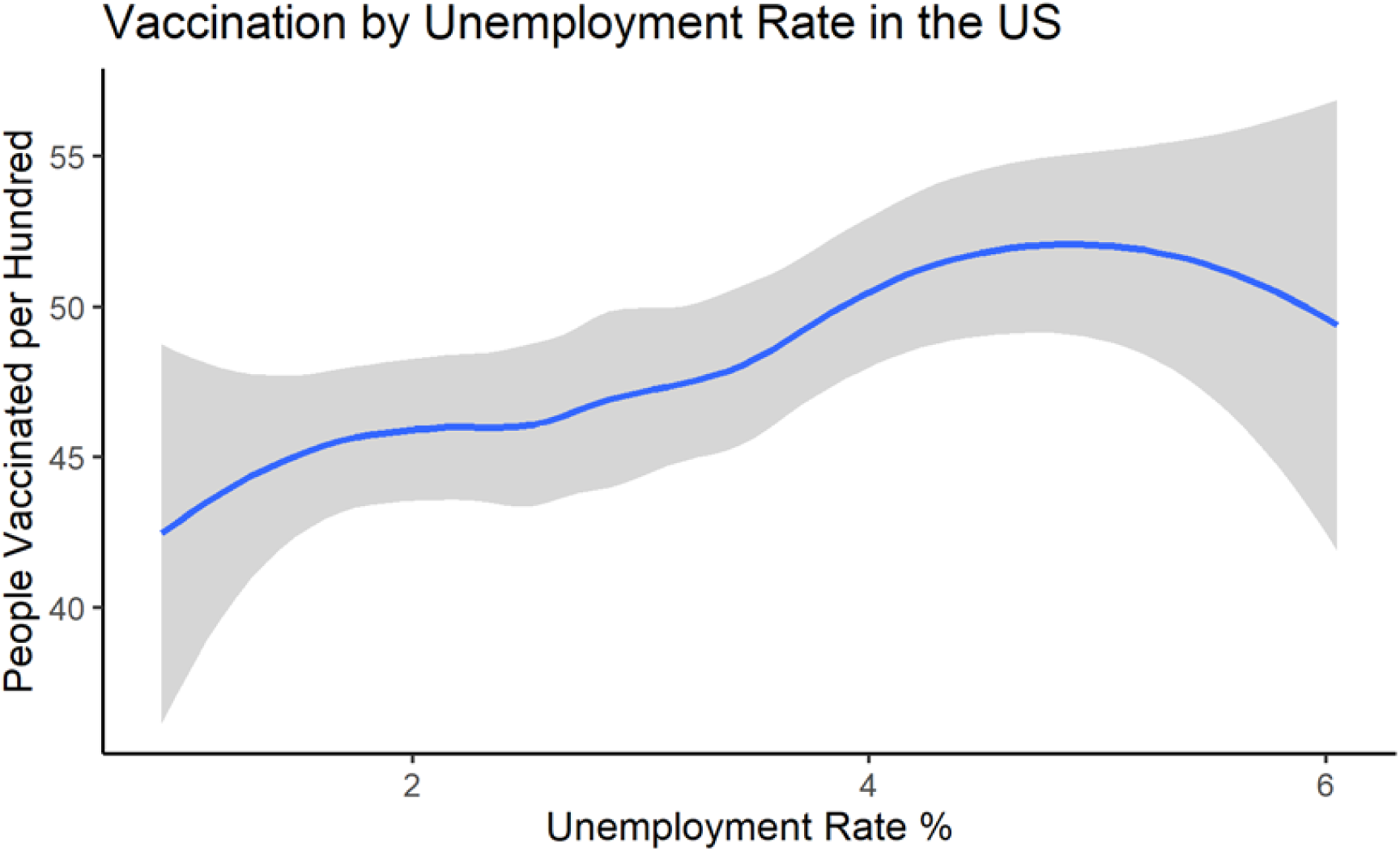

This study has some limitations. First, due to prioritizing a specific population for vaccination, the results might be influenced and prioritized for the elderly, occupational exposures, and people who have higher risks. Second, these associations are based on the overall information of each state, while individual-level data for each state could determine vulnerability in more detail.

## Conclusion

Our study indicates a positive relationship between higher vaccination rates and higher unemployment rates in the US at the state level. Although the regression analysis significantly shows this association, there is a contradiction in the states with higher than 5% unemployment, suggesting more research to understand this relationship. However, the current study reveals no association between most of the socioeconomic factors and vaccination coverage after the first 100 days of the vaccine initiation program. The high rate of vaccination among the unemployed population could be positive news on the economy, suggesting a decrease in disparity in the US. This finding represents a starting point for future research to look at individual-level data to determine vulnerability.

## Data Availability

Publicly available at https://ourworldindata.org/covid-vaccinations and https://github.com/stccenter/COVID-19-Data/tree/master/US.

## Data Availability

The data included in the study are publicly available.

https://ourworldindata.org/

https://www.covid19india.org/

## Code Availability

The code to perform all analyses described in Methods and Usage Notes sections was written in R-3.6.2 and is publicly available at https://ourworldindata.org/covid-vaccinations and https://github.com/stccenter/COVID-19-Data/tree/master/US.

## Author Approval

All authors have seen and approved the manuscript.

## Author Contributions

Ali Roghani designed the study and implemented methods and analyses. Samin Panahi contributed to evaluate and develop the methods, and structure. All authors were involved in developing the ideas and drafting the paper.

